# Short report: Ethnicity and COVID-19 death in the early part of the COVID-19 second wave in England: an analysis of OpenSAFELY data from 1^st^ September to 9^th^ November 2020

**DOI:** 10.1101/2021.02.02.21250989

**Authors:** Krishnan Bhaskaran, Rohini Mathur, Christopher T Rentsch, Caroline E Morton, William J Hulme, Anna Schultze, Brian MacKenna, Rosalind M Eggo, Angel YS Wong, Elizabeth J Williamson, Harriet Forbes, Kevin Wing, Helen I McDonald, Chris Bates, Seb Bacon, Alex J Walker, David Evans, Peter Inglesby, Amir Mehrkar, Helen J Curtis, Nicholas J DeVito, Richard Croker, Henry Drysdale, Jonathan Cockburn, John Parry, Frank Hester, Sam Harper, Ian J Douglas, Laurie Tomlinson, Stephen JW Evans, Richard Grieve, Liam Smeeth, Ben Goldacre

**Affiliations:** London School of Hygiene and Tropical Medicine, WC1E 7HT; The DataLab, Nuffield Department of Primary Care Health Sciences, University of Oxford, OX26GG; TPP, TPP House, 129 Low Lane, Horsforth, Leeds, LS18 5PX

**Author notes:** contributed equally (Krishnan Bhaskaran, Rohini Mathur). joint senior authors (Liam Smeeth, Ben Goldacre).

## Abstract

Black and minority ethnic groups were at raised risk of dying from COVID-19 during the first few months of the COVID-19 epidemic in England. We aimed to investigate whether ethnic inequalities in COVID-19 deaths were similar in the more recent “second wave” of the epidemic. Working on behalf of NHS England, we used primary care and linked ONS mortality data within the OpenSAFELY platform. All adults in the database at 1st September 2020 and with at least 1 year of prior follow-up and a record of ethnicity were included. The outcome was COVID-19-related death (death with COVID-19 listed as a cause of death on the death certificate). Follow-up was to 9th November 2020. Hazard ratios for ethnicity were calculated using Cox regression models adjusted for age and sex, and then further adjusted for deprivation. 13,223,154 people were included. During the study period, people of South Asian ethnicity were at higher risk of death due to COVID-19 than white people after adjusting for age and sex (HR = 3.47, 95% CI 2.99-4.03); the association attenuated somewhat on further adjustment for index of multiple deprivation (HR = 2.86, 2.46-3.33, Table 2). In contrast with the first wave of the epidemic, we found little evidence of a raised risk in black or other ethnic groups compared to white (HR for black vs white = 1.28, 0.87-1.88 adjusted for age and sex; and 1.01, 0.69-1.49 further adjusted for deprivation). Our findings suggest that ethnic inequalities in the risk of dying COVID-19-related death have changed between the first and early second wave of the epidemic in England.

## Background and aims

As reported by OpenSAFELY^1,2^ and others,^3^ black and minority ethnic groups were at raised risk of dying from COVID-19 during the first few months of the COVID-19 epidemic in England. We aimed to investigate whether ethnic inequalities in COVID-19 deaths were similar in the more recent “second wave” of the epidemic.

## Methods

The analysis was conducted within the OpenSAFELY platform on behalf of NHS England, using routinely collected electronic primary care data from practices using TPP SystmOne software, linked to ONS death registrations. The start date of the study was 1^st^ September 2020, chosen as being approximately the date of minimum COVID-19 mortality between the first and second waves of the epidemic in England. All adults under follow-up in a TPP general practice on 1^st^ September 2020, with at least one year of follow-up prior to this date, and with a record indicating their ethnicity were included. Ethnicity codes were classified into 5 main categories; a 16-level categorisation was used in a secondary analysis. COVID-19 mortality was defined as a record for death in linked ONS data (complete to 9^th^ November 2020) with the COVID related ICD-10 codes U071 or U072 listed as a cause of death anywhere on the death certificate. Following our previous methodology,^2^ Cox regression models were used to estimate the association between ethnicity and COVID-19 mortality, first adjusted for age (as a 4-knot cubic spline) and sex only, and then additionally adjusted for index of multiple deprivation, an area-based measure of socioeconomic status derived from the patient’s postcode. Analyses were stratified by the Sustainability and Transformation Partnership (STP, an NHS administrative region) of the patient’s general practice to account for geographical variation in infection rates across the country.

## Results

Of 17,695,074 people registered in TPP on 1^st^ September 2020 and with a year’s prior follow-up available, 13,223,154 (75%) had a record of ethnicity and were included (Table 1). People of South Asian ethnicity were at higher risk of death due to COVID-19 than white people during the study period after adjusting for age and sex (HR = 3.47, 95% CI 2.99-4.03); the association attenuated somewhat on further adjustment for index of multiple deprivation (HR = 2.86, 2.46-3.33, Table 2). Analysis using a finer categorisation of ethnicity suggested that the association was especially pronounced for Bangladeshi and Pakistani ethnicities. In contrast with results from the first wave of the epidemic, there was little evidence during the study period of raised risk in Black or Other ethnic groups compared to white (Figure 1). Further analysis stratified by age and sex showed similar patterns (Tables A1-A3). A further exploratory analysis suggested a largely consistent pattern across English regions (Table A4, Figure A1).

**Table 1:** Characteristics of the study population

|  | Ethnicity |  |  |  |  |
| --- | --- | --- | --- | --- | --- |
|  | White | Mixed | South Asian | Black | Other |
| N | 11,295,498 (100.0) | 179,253 (100.0) | 1,059,377 (100.0) | 355,195 (100.0) | 333,831 (100.0) |
| Age |  |  |  |  |  |
| 18-39 | 3,421,561 (30.3) | 97,323 (54.3) | 494,537 (46.7) | 147,586 (41.6) | 178,499 (53.5) |
| 40-49 | 1,838,647 (16.3) | 36,669 (20.5) | 252,690 (23.9) | 84,268 (23.7) | 72,072 (21.6) |
| 50-59 | 2,084,805 (18.5) | 25,606 (14.3) | 143,614 (13.6) | 70,052 (19.7) | 42,485 (12.7) |
| 60-69 | 1,720,122 (15.2) | 11,796 (6.6) | 96,704 (9.1) | 30,687 (8.6) | 23,887 (7.2) |
| 70-79 | 1,460,546 (12.9) | 5,202 (2.9) | 46,763 (4.4) | 12,899 (3.6) | 11,703 (3.5) |
| 80+ | 769,817 (6.8) | 2,657 (1.5) | 25,069 (2.4) | 9,703 (2.7) | 5,185 (1.6) |
| Median (IQR) | 51 (36-66) | 38 (28-50) | 41 (32-52) | 43 (32-54) | 38 (29-49) |
| Sex |  |  |  |  |  |
| Female | 5,872,445 (52.0) | 92,162 (51.4) | 508,560 (48.0) | 177,044 (49.8) | 163,852 (49.1) |
| Male | 5,423,053 (48.0) | 87,091 (48.6) | 550,817 (52.0) | 178,151 (50.2) | 169,979 (50.9) |
| Index of multiple deprivation quintile |  |  |  |  |  |
| 1 (least deprived) | 2,423,497 (21.5) | 23,885 (13.3) | 93,346 (8.8) | 24,089 (6.8) | 45,053 (13.5) |
| 2 | 2,378,336 (21.1) | 28,929 (16.1) | 121,274 (11.4) | 36,626 (10.3) | 57,590 (17.3) |
| 3 | 2,336,657 (20.7) | 34,831 (19.4) | 201,001 (19.0) | 59,057 (16.6) | 65,490 (19.6) |
| 4 | 2,174,386 (19.3) | 42,262 (23.6) | 304,523 (28.7) | 94,714 (26.7) | 81,802 (24.5) |
| 5 | 1,982,622 (17.6) | 49,346 (27.5) | 339,233 (32.0) | 140,709 (39.6) | 83,896 (25.1) |

**Table 2:** Association between ethnicity and COVID-19 death 1^st^ Sept - 9^th^ Nov 2020

|  | Ethnicity |  |  |  |  |
| --- | --- | --- | --- | --- | --- |
|  | White | Mixed | South Asian | Black | Other |
| COVID-19 deaths | 1771 | 8 | 224 | 27 | 16 |
| Person-years (millions) | 2,131,944 | 33,853 | 200,061 | 67,078 | 63,054 |
| Age-sex adjusted HR | 1.00 (REF) | 1.29 (0.64-2.58) | 3.47 (2.99-4.03) | 1.28 (0.87-1.88) | 1.26 (0.77-2.07) |
| + deprivation adjusted | 1.00 (REF) | 1.13 (0.56-2.26) | 2.86 (2.46-3.33) | 1.01 (0.69-1.49) | 1.14 (0.70-1.88) |
| Finer ethnic categories |  |  |  |  |  |
| Age-sex adjusted HRs | <u>British/Mixed</u><br><u>British</u><br>1.00 (REF)<br><u>Irish</u><br>0.84 (0.48-1.49)<br><u>Other white</u><br>1.09 (0.88-1.35) | <u>White+Black</u><br><u>Caribbean</u><br>0.85 (0.21-3.40)<br><u>White+Black African</u><br>2.05 (0.51-8.22)<br><u>White + Asian</u><br>0.93 (0.13-6.64)<br><u>Other mixed</u><br>1.62 (0.52-5.03) | <u>Indian/British Indian</u><br>2.70 (2.12-3.43)<br><u>Pakistani/British</u><br><u>Pakistani</u><br>4.43 (3.62-5.43)<br><u>Bangladeshi/British</u><br><u>Bangladeshi</u><br>5.96 (3.63-9.79)<br><u>Other Asian</u><br>2.49 (1.62-3.81) | <u>Caribbean</u><br>1.27 (0.79-2.07)<br><u>African</u><br>0.99 (0.41-2.38)<br><u>Other Black</u><br>1.68 (0.70-4.06) | <u>Chinese</u><br>1.07 (0.40-2.85)<br><u>Other</u><br>1.32 (0.75-2.34) |
| PREVIOUS RESULTS FOR COMPARISON<br>Study period 1st Feb to 6th May<br>(Williamson et al<sup>2</sup>) |  |  |  |  |  |
| Age-sex adjusted HRs | 1.00 (REF) | 1.62 (1.26-2.08) | 1.69 (1.54-1.84) | 1.88 (1.65-2.14) | 1.37 (1.13-1.65) |

**Figure 1:**
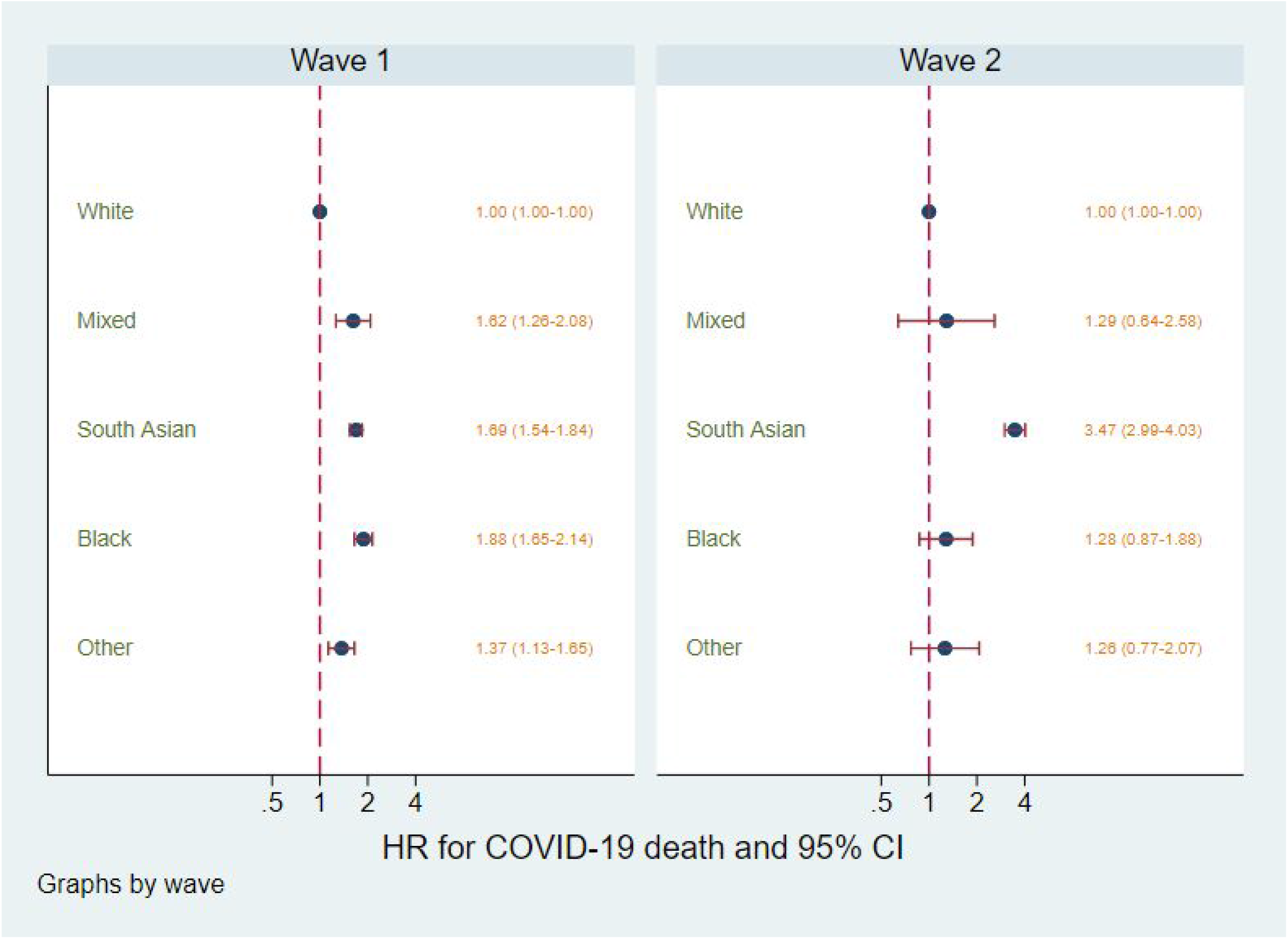
Association between ethnicity and COVID-19 death in wave 1 (1st Feb - 6th May)^2^ versus wave 2 (1^st^ Sept - 9^th^ Nov 2020), adjusted for age and sex

## Interpretation

Ethnic inequalities in the risk of dying COVID-19-related death appear to have changed between the first and early second wave of the epidemic in England. In the period between September and early November 2020, South Asian ethnic groups (in particular those of Bangladeshi and Pakistani ethnicity) were at markedly increased risk compared to people of white ethnicity, while people of Black and other ethnicities had similar risks to white. This suggests that mitigation strategies may need to be targeted towards South Asian communities.

Any underlying ethnic differences in the tendency to experience more severe disease once infected would be expected to remain relatively constant over time. Here we have observed marked changes in ethnic differences in outcomes within a short time period, strongly suggesting that ethnic inequalities in COVID-19 outcomes are driven by risk of infection as opposed to innate susceptibility to severe disease.

Ongoing monitoring of ethnic inequalities in COVID-19 outcomes, including by geographic region, is needed to detect any further changes in these patterns.

## Data Availability

All data were linked, stored and analysed securely within the OpenSAFELY platform https://opensafely.org/. All code is shared openly for review and re-use under MIT open license. Detailed pseudonymised patient data is potentially re-identifiable and therefore not shared. We rapidly delivered the OpenSAFELY data analysis platform without prior funding to deliver timely analyses on urgent research questions in the context of the global Covid-19 health emergency: now that the platform is established we are developing a formal process for external users to request access in collaboration with NHS England; details of this process will be published shortly on OpenSAFELY.org.

## Information governance and ethical approval

NHS England is the data controller; TPP is the data processor; and the key researchers on OpenSAFELY are acting on behalf of NHS England. This implementation of OpenSAFELY is hosted within the TPP environment which is accredited to the ISO 27001 information security standard and is NHS IG Toolkit compliant; patient data has been pseudonymised for analysis and linkage using industry standard cryptographic hashing techniques; all pseudonymised datasets transmitted for linkage onto OpenSAFELY are encrypted; access to the platform is via a virtual private network (VPN) connection, restricted to a small group of researchers; the researchers hold contracts with NHS England and only access the platform to initiate database queries and statistical models; all database activity is logged; only aggregate statistical outputs leave the platform environment following best practice for anonymisation of results such as statistical disclosure control for low cell counts. The OpenSAFELY research platform adheres to the data protection principles of the UK Data Protection Act 2018 and the EU General Data Protection Regulation (GDPR) 2016. In March 2020, the Secretary of State for Health and Social Care used powers under the UK Health Service (Control of Patient Information) Regulations 2002 (COPI) to require organisations to process confidential patient information for the purposes of protecting public health, providing healthcare services to the public and monitoring and managing the COVID-19 outbreak and incidents of exposure. Taken together, these provide the legal bases to link patient datasets on the OpenSAFELY platform. GP practices, from which the primary care data are obtained, are required to share relevant health information to support the public health response to the pandemic and have been informed of the OpenSAFELY analytics platform.

This study was approved by the Health Research Authority (REC reference 20/LO/0651) and by the LSHTM Ethics Board (reference 21863).

## SUPPLEMENTARY ANALYSIS

**Table A1:** HRs for the association between ethnicity and COVID-19 death stratified by sex, during the period 1^st^ Sept to 9^th^ Nov 2020

|  | Men | Women |
| --- | --- | --- |
| White | 1.00 (ref) | 1.00 (ref) |
| Mixed | 1.08 (0.41-2.90) | 1.59 (0.59-4.25) |
| South Asian | 4.02 (3.37-4.80) | 2.62 (2.03-3.40) |
| Black | 1.02 (0.59-1.78) | 1.66 (0.98-2.83) |
| Other | 1.31 (0.70-2.45) | 1.19 (0.53-2.65) |

From an age and sex adjusted model with interaction between ethnicity and sex. P-interaction = 0.04

**Table A2:** HRs for the association between ethnicity (in fine categories*) and COVID-19 death stratified by sex, during the period 1^st^ Sept to 9^th^ Nov 2020

|  | Men | Women |
| --- | --- | --- |
| **White groups** |  |  |
| White British | 1.00 (ref) | 1.00 (ref) |
| Other White | 1.10 (0.85-1.42) | 1.00 (0.74-1.36) |
| **Mixed** | 1.08 (0.40-2.89) | 1.57 (0.59-4.21) |
| **South Asian groups** |  |  |
| Indian | 3.33 (2.53-4.37) | 1.72 (1.10-2.71) |
| Pakistani | 4.94 (3.86-6.33) | 3.68 (2.61-5.20) |
| Bangladeshi | 7.25 (4.09-12.85) | 3.91 (1.46-10.48) |
| Other Asian | 2.74 (1.64-4.59) | 2.09 (0.99-4.42) |
| **Black groups** |  |  |
| Black Caribbean | 1.27 (0.68-2.38) | 1.29 (0.61-2.72) |
| Other Black | 0.61 (0.20-1.90) | 2.26 (1.07-4.77) |
| **Other** | 1.30 (0.70-2.44) | 1.17 (0.52-2.62) |

From an age and sex adjusted model with interaction between ethnicity and sex. \*All Mixed groups were combined, and Black African/Other Black were combined, due to small numbers of events. P-interaction = 0.15

**Table A3:** HRs for the association between ethnicity and COVID-19 death stratified by age, during the period 1^st^ Sept to 9^th^ Nov 2020

|  | Age <70 years | Age ≥70 years |
| --- | --- | --- |
| White | 1.00 (ref) | 1.00 (ref) |
| Mixed | 1.60 (0.51-5.02) | 1.19 (0.49-2.86) |
| South Asian | 4.65 (3.54-6.10) | 3.25 (2.73-3.87) |
| Black | 1.66 (0.82-3.36) | 1.21 (0.76-1.91) |
| Other | 0.88 (0.28-2.76) | 1.45 (0.84-2.51) |

From an age and sex adjusted model (age fitted as a spline as per original model), with interaction added between ethnicity and binary age group (<70/≥70). P-interaction = 0.18

**Table A4:** Region-specific observed and expected* deaths by ethic group

|  |  | White | Mixed | S Asian | Black | Other |
| --- | --- | --- | --- | --- | --- | --- |
| East | Population | **2695831** | **42128** | **169390** | **84801** | **61624** |
|  | Observed | 188 | <=5 | 12 | <=5 | 0 |
|  | Expected | - | 0.75 | 3.68 | 1.94 | 1.17 |
| E Midlands | Population | **2041696** | **29024** | **203169** | **58240** | **50336** |
|  | Observed | 357 | <=5 | 46 | <=5 | <=5 |
|  | Expected | - | 1.30 | 16.47 | 4.09 | 3.10 |
| London | Population | **572729** | **35563** | **250599** | **88759** | **92085** |
|  | Observed | 24 | 0 | 24 | 6 | 6 |
|  | Expected | - | 0.88 | 8.15 | 3.33 | 2.07 |
| North East | Population | **565126** | **6366** | **28299** | **11041** | **13750** |
|  | Observed | 216 | <=5 | 11 | 0 | <=5 |
|  | Expected | - | 0.51 | 3.86 | 1.68 | 1.13 |
| North West | Population | **1113418** | **6428** | **39869** | **9145** | **16323** |
|  | Observed | 354 | <=5 | 21 | 0 | <=5 |
|  | Expected | - | 0.36 | 4.02 | 0.54 | 1.16 |
| South East | Population | **716772** | **11697** | **32274** | **13276** | **22861** |
|  | Observed | 42 | 0 | <=5 | 0 | <=5 |
|  | Expected | - | 0.15 | 0.54 | 0.16 | 0.31 |
| South West | Population | **1588697** | **14890** | **37453** | **12999** | **23906** |
|  | Observed | 81 | 0 | <=5 | 0 | <=5 |
|  | Expected | - | 0.18 | 0.49 | 0.21 | 0.30 |
| W Midlands | Population | **379409** | **13013** | **104530** | **40173** | **18479** |
|  | Observed | 90 | <=5 | 28 | 11 | <=5 |
|  | Expected | - | 0.79 | 9.96 | 5.30 | 1.23 |
| Yorkshire & Humber | Population | **1620066** | **20028** | **193702** | **36544** | **34428** |
|  | Observed | 418 | <=5 | 77 | <=5 | <=5 |
|  | Expected | - | 1.30 | 18.32 | 3.62 | 2.00 |

Non-zero counts of <=5 are redacted. Observed = observed COVID-19 deaths. \*Expected = expected COVID-19 deaths if the age, sex and region-specific COVID-19 death rates for the local White population applied (for this analysis, age was grouped as 18-39, 40-49, 50-59, 60-69, 70-79, 80+).

**Figure A1:**
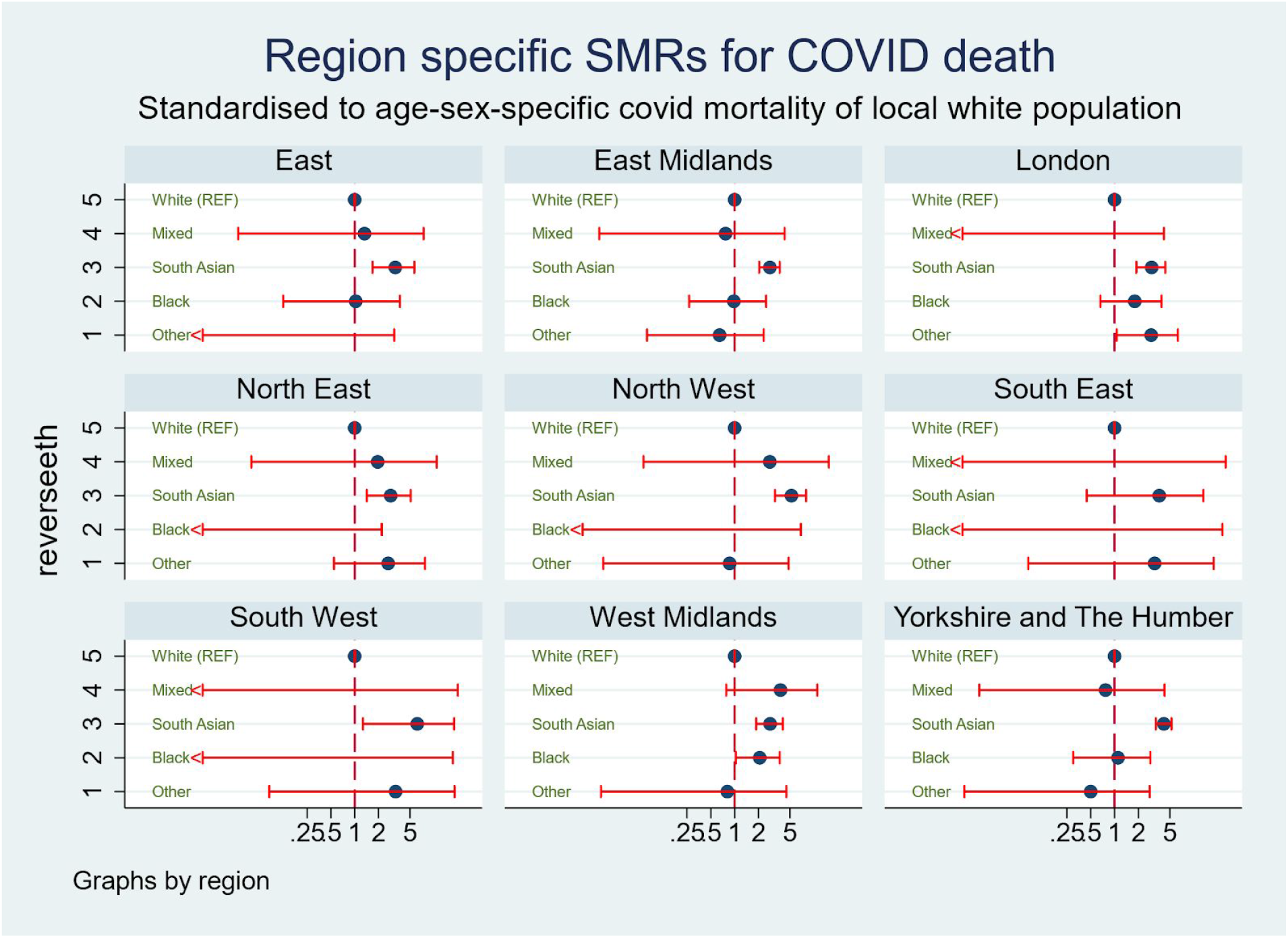
Region-specific Standardised Mortality Ratios (SMR) for COVID-19 death SMRs calculated for each region using White (in that region) as the reference; SMRs are Observed/Expected where Expected = expected COVID-19 deaths if the age, sex and region-specific COVID-19 death rates for the local White population applied (for this analysis, age was grouped as 18-39, 40-49, 50-59, 60-69, 70-79, 80+).

